# Detecting Glaucoma Worsening Using Optical Coherence Tomography Derived Visual Field Estimates

**DOI:** 10.1101/2024.10.17.24315710

**Authors:** Alex T. Pham, Chris Bradley, Kaihua Hou, Patrick Herbert, Mathias Unberath, Pradeep Y. Ramulu, Jithin Yohannan

**Author notes:** **Corresponding author:** Jithin Yohannan, MD MPH Wilmer Eye Institute, Johns Hopkins Hospital 600 N. Wolfe St., Baltimore, MD 21287, USA. **Meeting Presentation:** Association for Research in Vision and Ophthalmology (ARVO) 2023. **Reprint Address:** Wilmer Eye Institute, Johns Hopkins Hospital, 600 N. Wolfe St., Baltimore, MD 21287, USA.

## Abstract

**Objective:** Multiple studies have attempted to generate visual field (VF) mean deviation (MD) estimates using cross-sectional optical coherence tomography (OCT) data. However, whether such models offer any value in detecting longitudinal VF progression is unclear. We address this by developing a machine learning (ML) model to convert OCT data to MD and assessing its ability to detect longitudinal worsening.

**Design:** Retrospective, longitudinal study

**Participants:** A model dataset of 70,575 paired OCT/VFs to train an ML model converting OCT to VF-MD. A separate progression dataset of 4,044 eyes with ≥ 5 paired OCT/VFs to assess the ability of OCT-derived MD to detect worsening. Progression dataset eyes had two additional unpaired VFs (≥ 7 total) to establish a “ground truth” rate of progression defined by MD slope.

**Methods:** We trained an ML model using paired VF/OCT data to estimate MD measurements for each OCT scan (OCT-MD). We used this ML model to generate longitudinal OCT-MD estimates for progression dataset eyes. We calculated MD slopes after substituting/supplementing VF-MD with OCT-MD and measured the ability to detect progression. We labeled true progressors using a ground truth MD slope <0.5 dB/year calculated from ≥ 7 VF-MD measurements. We compared the area under the curve (AUC) of MD slopes calculated using both VF-MD (with <7 measurements) and OCT-MD. Because we found OCT-MD substitution had a statistically inferior AUC to VF-MD, we simulated the effect of reducing OCT-MD mean absolute error (MAE) on the ability to detect worsening.

**Main Outcome Measures:** AUC

**Results:** OCT-MD estimates had an MAE of 1.62 dB. AUC of MD slopes with partial OCT-MD substitution was significantly worse than the VF-MD slope. Supplementing VF-MD with OCT-MD also did not improve AUC, regardless of MAE. OCT-MD estimates needed an MAE ≤ 1.00 dB before AUC was statistically similar to VF-MD alone.

**Conclusion:** ML models converting OCT data to VF-MD with error levels lower than published in prior work (MAE: 1.62 dB) were inferior to VF-MD data for detecting trend-based VF progression. Models converting OCT data to VF-MD must achieve better prediction errors (MAE ≤ 1 dB) to be clinically valuable at detecting VF worsening.

## Introduction

Early detection of glaucoma progression is critical to manage the disease effectively. Identifying those at the highest risk of progression allows clinicians to adjust therapy before additional irreversible vision loss occurs. Glaucoma monitoring is usually done by tracking structural changes with optical coherence tomography (OCT) imaging and functional changes with visual field (VF) testing. Since OCT imaging and VF testing have their respective advantages and disadvantages that make either alone less than ideal for detecting progression, they are often used in combination. In general, there are differences in the ability of VF and OCT to detect glaucoma worsening at various stages of the disease. OCT is more sensitive to detecting disease progression in earlier stages of glaucoma. At the same time, VF is more informative at later stages when structural features reach the measurement floor of the OCT instrument.^1–7^

There are multiple approaches to addressing the difference between OCT and VF to monitor progression. While clinicians are skilled at monitoring progression using their own experience and judgment, it has been suggested that OCT could help guide VF testing by focusing on measuring functional changes in regions with significant structural changes as the structure-function relationship demonstrates better agreement with regional or sectoral measurements.^8^ Another approach involves using structural information from OCT, such as the retinal nerve fiber layer (RNFL), to predict functional measures, such as mean deviation (MD).^9–16^ The appeal of this pursuit is that it relates different measurement scales (microns per year versus dB per year), and it provides a “functional” measure while retaining the inherent clinical advantages of OCT, such as better repeatability, reproducibility, objectivity, and sensitivity to early glaucomatous damage.^3–5,17–20^

Due to the growing availability of large datasets from electronic health records, most recent efforts to predict MD from OCT (OCT-MD) have focused on applying deep learning models to optic nerve head OCT scans, macular OCT scans, or both.^9–14,16^ However, the OCT-MD estimations from these models have limited accuracy, with mean absolute errors (MAE) ranging from 2-5 dB ^9–14,16^The test-retest variability of MD measurements from VF (VF-MD) is less than 1.5 dB.^21,22^ The best-performing structure-function models in the work mentioned above report MAEs of approximately 2 dB. It is likely that OCT-MD alone cannot predict progression with the same accuracy as VF, but this has yet to be assessed. Additionally, OCT-MD estimates could still have clinical utility if assessed in combination with VF measurements. This is relevant because patients often alternate between OCT and VF and frequently have both available for the clinician to assess progression. Assuming at least five VFs are needed to calculate a reliable MD rate of change for monitoring trend-based progression, the ability to substitute VF-MD with OCT-MD would reduce the testing burden on patients and allow clinicians to determine the rate of change more quickly, leading to earlier detection of progression.^23,24^ This is especially important since treatment decisions must often be made only after a few visits.

Hence, the aims of our study are two-fold. First, our study aimed to evaluate whether OCT-MD has any clinical value as a substitute for VF-MD (either through complete or partial substitution) to detect trend-based glaucoma progression with non-inferior accuracy to VF-MD alone. Second, if we find that OCT-MD estimates, with an MAE similar to or better than prior published work, are not accurate enough to be clinically viable, we aimed to determine the MAE needed for OCT-MD to be useful in trend-based analysis. Currently, there is no established evidence-based MAE threshold that investigators developing these models should aim to stay below. Thus, knowing the maximum acceptable error level would clarify the ideal goal for modeling the structure-function relationship to detect functional change over time.

## Methods

Our study adhered to the Declaration of Helsinki and was approved by the Johns Hopkins University School of Medicine Institutional Review Board.

### Study Population and Data Collection

Adult patients with a glaucoma or glaucoma-related diagnosis followed at the Wilmer Eye Institute Glaucoma Center of Excellence from April 2013 to July 2022 were considered eligible for our study. From these eligible eyes, we created a dataset to train and test a machine learning model to estimate OCT-MD, referred to as the “model dataset”, and a separate dataset to evaluate the ability to predict VF worsening from trend-based analysis of the OCT-MD generated by the model, referred to as the “progression dataset”. The same eye or patient was never present in both datasets. For the progression dataset, the inclusion criteria were eyes with 5 or more reliable VFs (reliability criteria defined below), each paired with a reliable optic nerve head OCT scan (reliability criteria defined below) taken within a 1-year time window. Each pairing was unique in that there were no overlapping VFs or OCTs among the pairings. These eyes also had to have 2 additional unpaired VF tests. The rationale for these inclusion criteria was based on the notion that at least 5 VF tests are needed to calculate a reliable MD slope^23,24^, and the additional 2 unpaired VF tests were used to establish a ground truth for the rate of progression. Hence, each eye in the progression dataset had 7 or more VFs. For the model dataset, the inclusion criteria were peripapillary OCT scans and VF tests obtained from eyes with less than 7 VFs over time, so there are no overlapping eyes between the progression and model dataset. OCT scans and VF tests also had to be paired within one year of each other. Only reliable OCT scans and reliable VF tests were considered during the construction of the model dataset as well.

For OCT scans to be considered reliable, they had to have a signal strength > 6, and average, superior quadrant, and inferior quadrant RNFL thickness measurements between 57 to 135, 175, and 190 μm, respectively. An RNFL floor of 57 μm was used because values below this threshold are likely due to artifact or segmentation error.^25,26^ Moreover, it is unlikely any further longitudinal changes can be observed in eyes that have reached the OCT floor.^27^ An RNFL ceiling of 135, 175, and 190 μm was used for the average, superior, and inferior thickness, respectively, because these thresholds are approximately three standard deviations above the average RNFL thickness measured in normal healthy eyes.^28^ All OCT studies were obtained using CIRRUS HD-OCT (Zeiss, Dublin, CA).

To be considered reliable, VFs had to have false positive rates < 15% for all stages of disease, false negative rates < 25% for suspect or mild glaucoma, and false negative rates < 50% for advanced glaucoma.^29^ All VF tests were performed using the Humphrey Visual Field Analyzer II or III with the SITA Standard, Fast, or Faster testing algorithm and the 24-2 pattern. Only VF tests with MD measurements better than –10.4 dB were included in the study because MDs worse than –10.4 dB likely indicate that the eye has reached the RNFL floor.^27^ Since our study aims to assess the feasibility of detecting progression with OCT-derived MD, the rationale for this restriction is to provide the most favorable conditions that would produce the most accurate conversion estimates. If the progression cannot be accurately detected using estimates limited to the dynamic range of OCT, it is unlikely to have the potential for clinical utility in real-world circumstances.

Variables collected for our study included age, gender, race, and glaucoma severity. Glaucoma severity was determined using the MD measurement from the first VF available for each eye. Eyes with a baseline MD better than −6 dB and between −6 and −10.4 dB were considered suspect/mild and moderate glaucoma, respectively. Eyes with an MD better than −6 dB were considered glaucoma suspect, as opposed to mild glaucoma, if their glaucoma hemifield test was “within normal limits”.

### Estimating VF Mean Deviation from Optic Nerve OCT Characteristics in the Model Dataset

Using the model dataset, we investigated multiple machine-learning models to estimate MD based on various features measured by OCT scans of the optic disc. Input features for each model were the average RNFL thickness, four quadrant RNFL thicknesses (superior, inferior, nasal, temporal), 12 clock hour RNFL thicknesses, 6 Garway-Heath Zone RNFL thicknesses, cup volume, disc area, rim area, average cup-to-disc ratio, vertical cup-to-disc ratio, signal strength, and baseline age. Classical machine learning algorithms that were tested included random forest, support vector machine (SVM) regression, lasso regression, and K-nearest neighbors. We also tested deep learning models such as convolutional neural networks, multi-layer perceptron, and a model combining both previous neural networks. A training and testing set were created using an 80:20 percent split of the model dataset described above. Hyperparameters were optimized by comparing the performance of various combinations of parameters after cross-validation on the training set. We used 5-fold cross-validation, which involves randomly splitting the training set into five non-overlapping folds of equal size. The model was then trained on a combination of four folds while the remaining fold, a holdout set, was used for validation to evaluate performance. An evaluation score was obtained from this holdout set, and this process was repeated five times so that each fold had an opportunity to be used once for validation. The model’s performance for a particular combination of hyperparameters was then summarized by taking the average of the evaluation scores from the five iterations. The combination of parameters that produced the strongest average evaluation score after cross-validation was used as the optimal parameters for the model. Afterward, the performance of each optimized model was evaluated by generating an OCT-MD from each paired OCT scan in the test set and calculating the mean absolute error (MAE) between the OCT-MD estimate and the real MD measurement, VF-MD. The optimized model with the lowest MAE obtained from the test set was used to predict disease progression described in the following section.

### Detecting Glaucoma Progression in the Progression Dataset

An overview of our study method can be seen in Figure 1. After constructing a model to generate OCT-MD, an MD estimate was obtained from each paired OCT scan for each eye in the progression dataset. We evaluated the utility of using these OCT-MD estimates with VF-MD to detect trend-based glaucoma progression. Our primary approach involved selecting a random subset of the longitudinal VF-MD measurements, replacing them with their paired OCT-MD estimates, and calculating the MD slope using ordinary least squares regression. We refer to this approach of combining OCT-MD and VF-MD as ‘substitution’. The MD slope calculation from substitution is referred to as the ‘VF-MD/OCT-MD slope’. Varying amounts of VF-MD substitutions were tested: 20%, 40%, 60%, 80%, and 100% substitution. For example, if an eye had ten VF studies with ten paired OCT scans, 80% substitution means that randomly chosen eight VF-MDs were replaced with their paired OCT-MDs.

**Figure 1:**
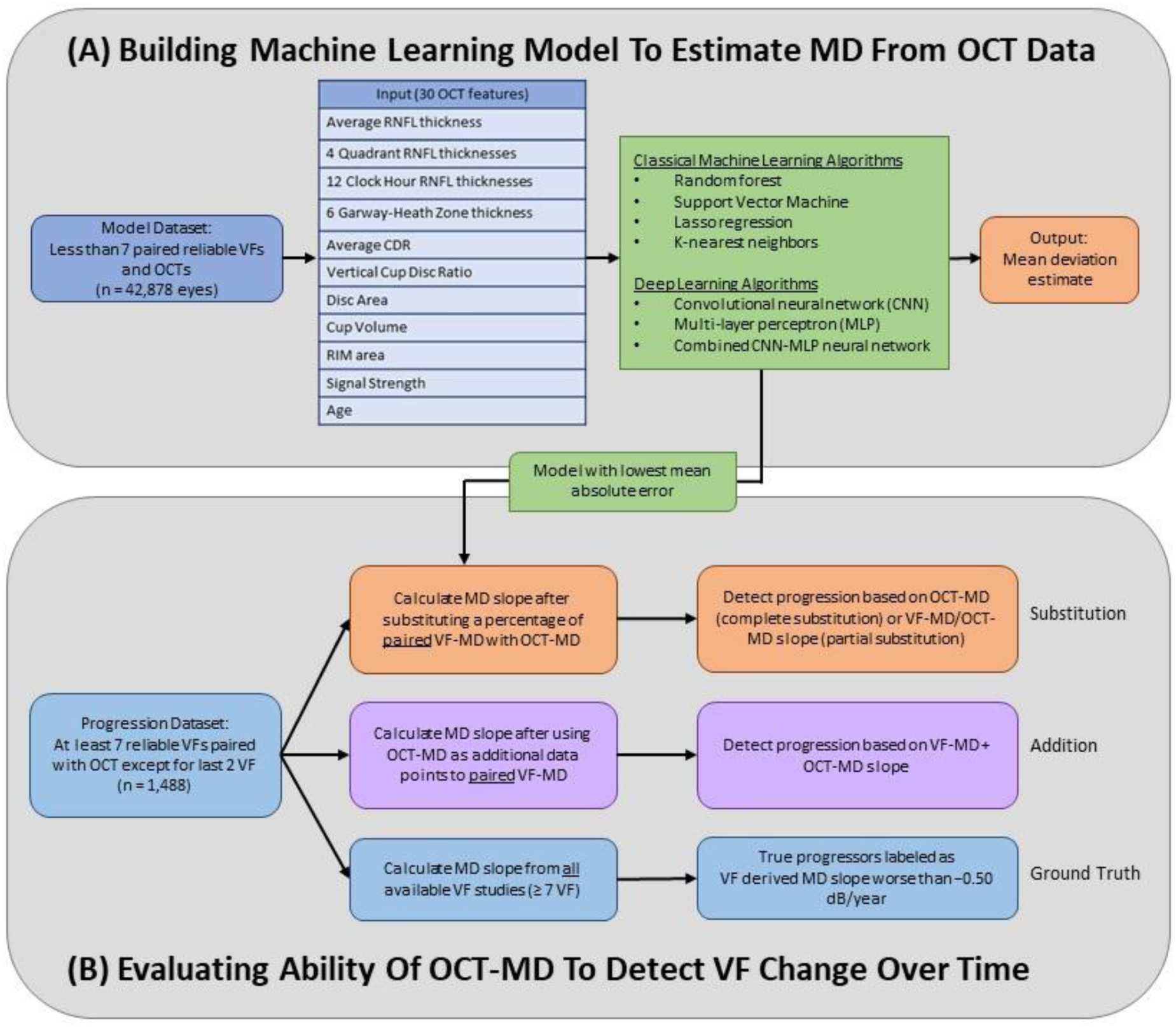
(A) Various models were built using data obtained from OCT scans to estimate a corresponding MD. (B) We then compared the ability to detect glaucoma progression using OCT-MD estimates with VF-MD measurements to using only VF-MD alone. To establish a ground truth for both, eyes were labeled true progressors if the MD slope calculated from all available VF studies (equating to at least 2 or more additional data points) was worse than −0.50 dB/year.

We were also interested in investigating whether OCT-MD could be used to improve the predictive ability of VF-MD when it is included as supplemental information in trend-based analysis. In other words, our secondary approach involved combining all OCT- MD estimates with all paired VF-MD measurements by using OCT-MD as additional data points in the MD slope calculation with ordinary least squares regression. We refer to this approach of combining OCT-MD and VF-MD as ‘addition’. The MD slope calculation from addition is referred to as the ‘VF-MD + OCT-MD slope’.

To evaluate the accuracy of the VF-MD/OCT-MD slopes and VF-MD + OCT-MD slopes, we established a “ground truth” MD slope for analysis purposes. The ground truth MD slope was calculated using all available VF studies (at least 7 or more), which equate to two additional data points beyond the time window of the paired VF tests and OCT scans (at least 5 or more) for each eye in the progression dataset. Receiver operating characteristic curves were generated for VF-MD/OCT-MD slopes and VF-MD + OCT- MD slopes using the ground truth MD slope. Eyes with a statistically significant ground truth MD slope (α = 0.05) worse than −0.50 dB/year were labeled true progressors.^30–33^ We also repeated the analysis to determine whether OCT-MD could predict slower and faster rates of VF worsening by using MD slope cut-offs of −0.25, −0.75, and −1.00 dB/year. The area under the receiver operating characteristic curve (AUC) was used to evaluate the performance of the VF-MD/OCT-MD and VF-MD + OCT-MD slopes.

To compare the VF-MD/OCT-MD slopes derived from substitution to VF-MD, we calculated an MD slope when no VF-MDs were substituted, which we will refer to as the baseline VF-MD slope. The VF-MD/OCT-MD slopes for 20%, 40%, and 60% substitution were also compared to baseline VF-MD slopes calculated using 20%, 40%, and 60% fewer VFs to determine whether partial substitution with OCT-MD does better than simply using fewer VFs to calculate MD slope. Statistical comparisons between the AUCs of the MD slopes were made using Delong’s test.

### Accuracy Needed to Predict Progression with OCT- MD

After evaluating the performance of VF-MD/OCT-MD and VF-MD + OCT-MD slopes in predicting progression, we investigated the impact of OCT-MD MAE on the AUCs. It is currently unknown what MAE is needed for OCT-MD estimates to be viable when used alone or combined with VF-MD in trend-based analysis to detect progression. To simulate OCT-MD model estimates with a lower MAE, we took the original MAE of our model and calculated the error percent reduction needed to lower it to 1.50, 1.25, 1.00, 0.75, and 0.50 dB. Then, we calculated the residual error between the paired OCT-MD and VF-MD for the eyes in our progression dataset. The OCT-MD estimates were brought closer to the paired VF-MD measurements by artificially reducing the residual error between them by the percentages calculated above. As an example, if the original MAE of our model was 2.00 dB, the percent reduction needed to achieve an MAE of 1.50 dB would be 25%. If the original OCT-MD estimate was −2.00 dB and the paired VF-MD was −1.00 dB, the residual error would be 1.00 dB. A 25% reduction of the residual would lead to a simulated OCT-MD estimate of −1.75 dB. MD slopes were recalculated using these simulated OCT-MD estimates, and the AUC analysis was repeated.

### Sensitivity Analyses

We conducted two sensitivity analyses. First, since our current inclusion criteria allow OCT scans and VFs to be paired up to one year apart, this may introduce temporal bias in the model. The OCT-MD produced by the model could be estimating a VF-MD one year ahead or behind the OCT scan date, which may affect our analysis of longitudinal changes to predict progression. To address this concern, we conducted a sensitivity analysis by training our machine learning model on only OCT scans and VF tests that were paired on the same day. In addition, when using the OCT-MD estimates to detect glaucoma progression, we only analyzed eyes with 5 or more OCT scans and VF tests also paired on the same day. Second, we investigated including confidence intervals when labeling true progressors since OCT-MD may not be able to detect progression in those with nosier VF tests. The confidence interval for a statistically significant MD slope had to be within a 0.50 dB range (+/- 0.25 dB of the slope) to be considered as a true progressor.

## Results

Baseline demographics, VF, and OCT characteristics for the model and progression datasets are shown in Table 1. The model dataset consisted of 70,575 paired optic disc OCT scans and VF studies obtained from 44,659 eyes, each with less than 7 reliable VFs. The progression dataset consisted of 4,044 eyes with at least 7 reliable VFs, with all but the last 2 VFs paired with OCT. The mean (SD) duration of time between OCT scans and VF studies for each pair was 102 (117) days. The mean age was slightly older in the progression dataset than in the model dataset (64 vs. 62 years, p < 0.001). Race, gender, and baseline glaucoma severity were similar for both datasets. Mean MD was slightly worse in the model dataset than in the progression dataset (–1.86 vs. –1.59 dB, p < 0.001). Pattern standard deviation was slightly better in the model dataset than in the progression dataset (2.46 vs. 2.56, p = 0.001). RNFL was slightly thinner (83 vs. 86 µm, p < 0.001) and CDR slightly larger (0.62 vs. 0.60, p < 0.001) in the progression dataset than the model dataset.

**Table 1:** Baseline demographic, VF, OCT characteristics.

|  | **Model dataset**<br>**(n = 44,659)** | **Progression dataset**<br>**(n=4,044)** |
| --- | --- | --- |
| **Mean age (SD), years** | 62 (15) | 64 (11) |
| **Gender** |  |  |
| Male, n | 18,860 (42%) | 1,715 (42%) |
| Female, n | 25,794 (58%) | 2,329 (58%) |
| **Race** |  |  |
| White or Caucasian, n | 24,413 (58%) | 2,305 (57%) |
| Black or African American, n | 13,068 (29%) | 1,360 (34%) |
| Asian, n | 3,220 (7.2%) | 196 (4.9%) |
| American Indian or Alaska Native, n | 150 (0.34%) | 9 (0.22%) |
| Pacific Islander, n | 48 (0.11%) | 4 (0.10%) |
| Hispanic, n | 2 (0.00%) | 0 (0%) |
| Other, n | 2,643 (5.9%) | 125 (3.1%) |
| Unknown, n | 1,115 (2.5%) | 45 (1.1%) |
| **Glaucoma Severity** |  |  |
| Suspect, n | 15,000 (34%) | 2,065 (51%) |
| Mild, n | 25,984 (58%) | 1,796 (44%) |
| Moderate, n | 3,675 (8.2%) | 183 (4.5%) |
| **VF Characteristics** |  |  |
| Mean MD (SD), dB | -1.86 (2.54) | -1.59 (2.22) |
| Mean PSD (SD), dB | 2.56 (1.89) | 2.46 (1.72) |
| **OCT Characteristics** |  |  |
| Mean RNFL (SD), μm | 86 (12) | 83 (11) |
| Mean CDR (SD) | 0.60 (0.16) | 0.62 (0.15) |

\* SD = standard deviation, MD = mean deviation, PSD = pattern standard deviation, RNFL = retinal nerve fiber thickness layer, CDR = cup-to-disc-ratio, VF = visual field, OCT = optical coherence tomography

Among the different machine learning models evaluated, the SVM model had the lowest MAE and was used for the remainder of the study. Table 2 shows the MAE of the SVM model and the percentage of OCT-MD estimates within 0.25 dB, 0.50 dB, 1.00 dB, 2.00 dB, and 4.00 dB of the true MD value. Overall, the MAE was 1.62 dB, and the percentage of estimates with 0.25 dB, 0.5 dB, 1 dB, 2 dB, and 4 dB of error were 11%, 21%, 41%, 71%, and 93%, respectively. The estimations became more inaccurate for later stages of disease. The estimates from eyes with suspect and mild glaucoma had an MAE below 2 dB and the highest proportion of estimates within the various margins of error. Estimates from eyes with moderate disease had the worst MAE of 5.55 dB and the lowest proportions within the various margins of error. This trend is also seen in Figure 2, demonstrating that the MAE of OCT-MD estimates increases the further VF-MD is from –1.0 dB.

**Figure 2:**
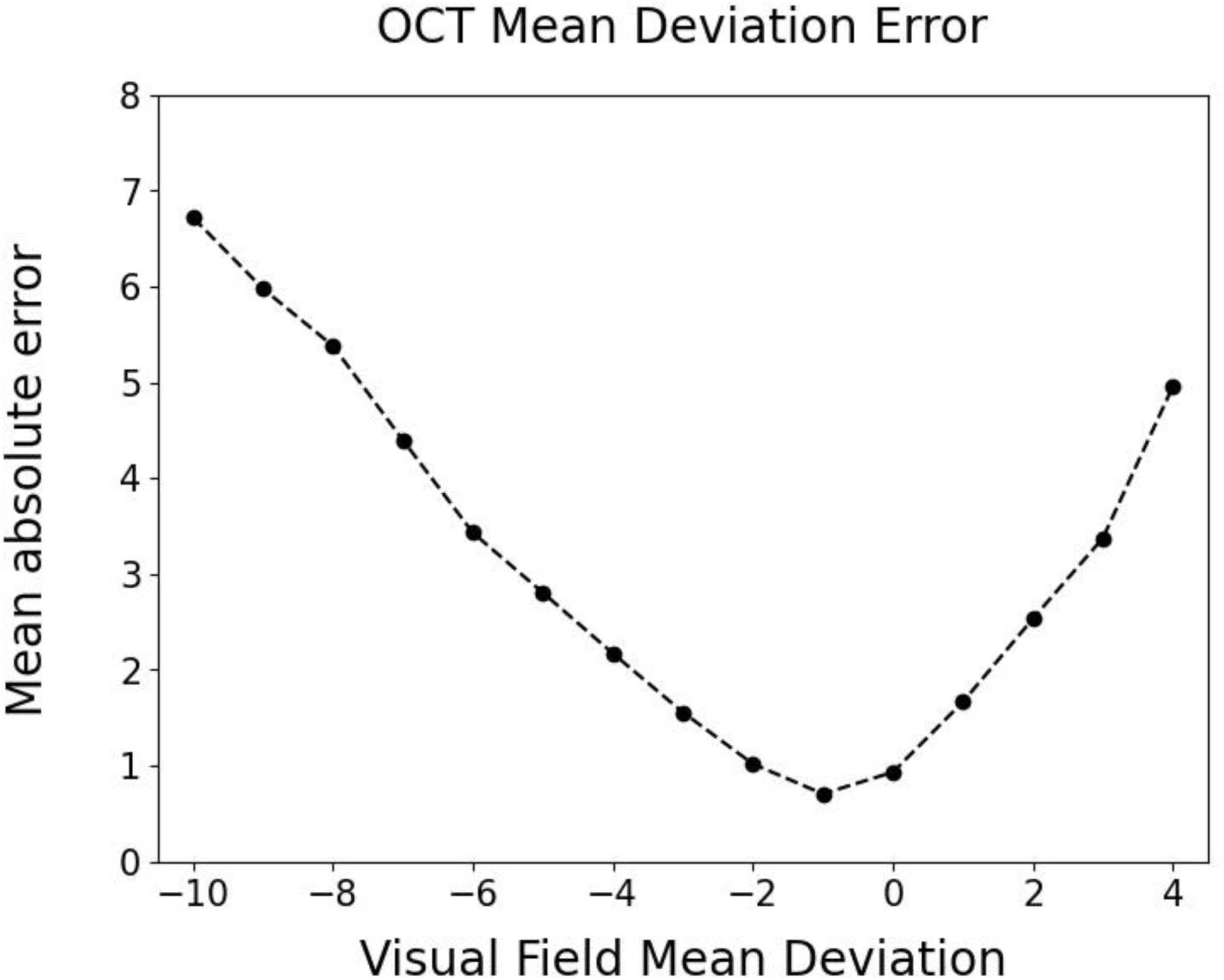
Mean absolute error of OCT-MD estimates across a range of VF-MD measurements

**Table 2:** OCT-MD Support Vector Machine regression performance metrics.

|  | Mean absolute error (dB) | % within 0.25 dB | % within 0.5 dB | % within 1 dB | % within 2 dB | % within 4 dB |
| --- | --- | --- | --- | --- | --- | --- |
| Overall (n = 12,222) | 1.62 | 11% | 21% | 41% | 71% | 93% |
| Suspect (n = 5,973) | 1.36 | 12% | 24% | 44% | 77% | 98% |
| Mild (n = 5,337) | 1.48 | 11% | 21% | 43% | 72% | 95% |
| Moderate (n = 912) | 5.55 | 1.5 % | 2.4% | 3.9% | 8.2% | 21% |

\* OCT-MD = OCT derived mean deviation estimates; n = the number of OCT-VF pairs from each dataset

### Ability of MD slope to detect progression when combining VF-MD with OCT-MD

Figure 3 demonstrates the diagnostic ability of VF-MD/OCT-MD slopes (substitution) calculated from various substitution percentages. For an MD slope cutoff of 0.50 dB/year, the AUCs (95% CI) for 20%, 40%, 60%, 80%, and 100% substitution are 0.88 (0.86 to 0.90), 0.84 (0.81 to 0.86), 0.77 (0.74 to 0.80), 0.70 (0.67 to 0.74), and 0.60 (0.58 to 0.64), respectively. The AUC (95% CI) of the baseline VF-MD slope is 0.91 (0.90 to 0.93) and corresponds to 0% substitution. The AUC of the baseline VF-MD slopes calculated using 20%, 40%, and 60% fewer VFs is 0.90 (0.88 to 0.92), 0.86 (0.83 to 0.88), and 0.77 (0.73 to 0.80). The AUC of 40% to 100% substitution was significantly worse than the baseline VF-MD slope. Although the AUC of 20% substitution was statistically similar to the baseline VF-MD slope with 0% substitution, it was also similar to the AUC of baseline VF-MD slopes calculated using 20% fewer VFs. The AUC of the OCT-MD slope from 40% and 60% substitution was similar to the baseline VF-MD slope calculated using 40% and 60% fewer VFs, respectively, as well. Figure 3 also shows the AUCs for substitution when using faster and slower MD slope cut-offs. The number of progressing eyes using an MD slope cutoff of –0.25, –0.50, –0.75, and –1.00 dB/year were 380, 149, 47, and 19 eyes, respectively.

**Figure 3:**
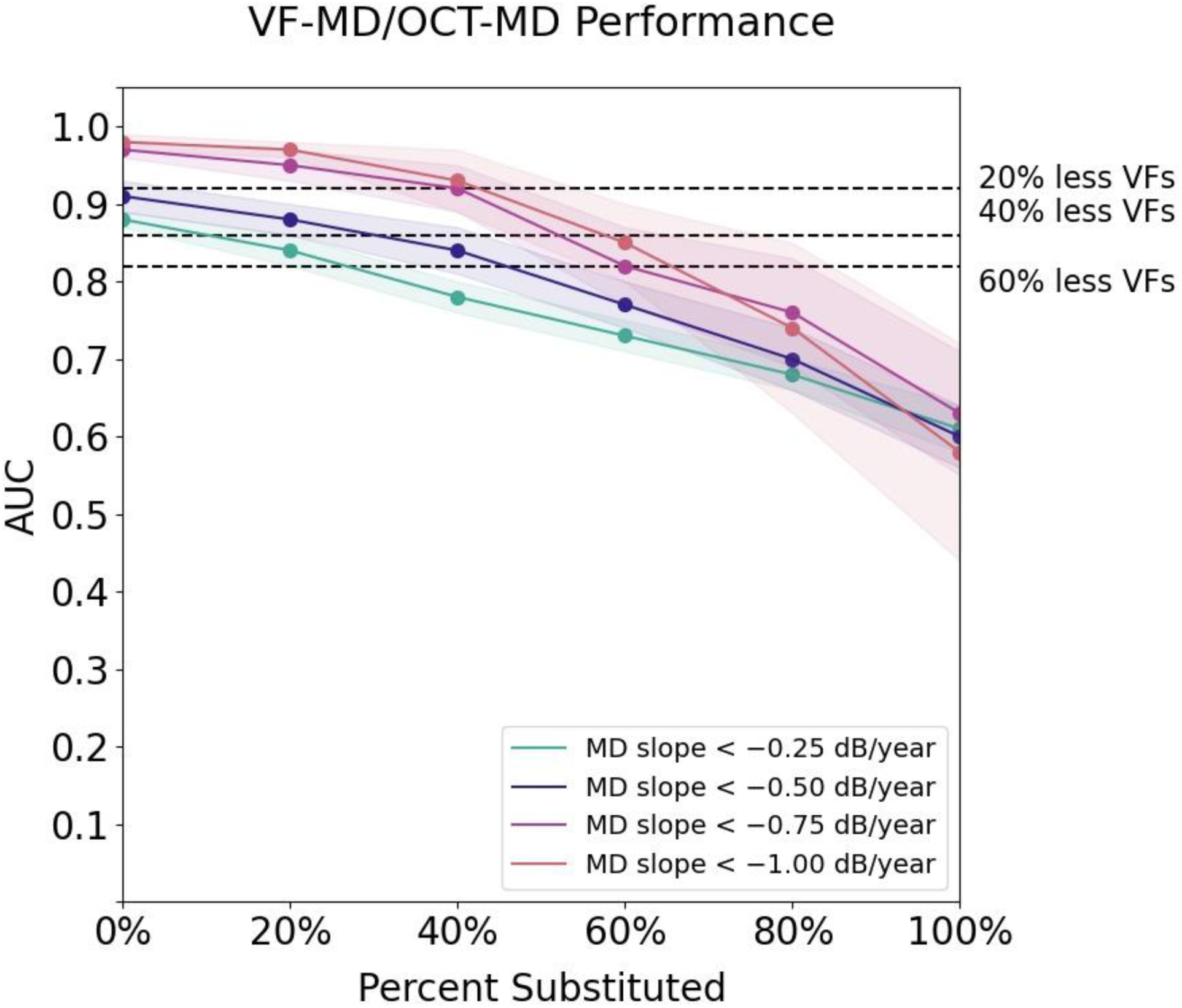
The predictive ability of VF-MD/OCT-MD slope calculated with different percentages substitution for various MD-slope cutoff thresholds. Substitution of 0% represents the performance of MD slopes calculated only from VF-MD (baseline VF-MD slope). A substitution of 100% represents the performance of MD slopes calculated from only OCT-MD. Dashed lines represent the performance of baseline VF-MD slopes using 20%, 40%, and 60% fewer VFs.

Table 3 shows the AUCs of the VF-MD + OCT-MD slope (addition) compared to the baseline VF-MD slope for various MD slope cutoff thresholds. For an MD slope cutoff of 0.50 dB/year, the AUC when using OCT-MD as additional data points is 0.89 (0.87 to 0.91) and was statistically similar to the baseline VF-MD slope. VF-MD + OCT-MD slopes were also statistically similar to the baseline VF-MD slope for slower and faster MD-slope cutoffs.

### Accuracy Simulation

The diagnostic ability of VF-MD/OCT-MD slopes with an MAE of 1.50, 1.25, 1.00, 0.75, and 0.50 dB is shown in Figure 4. Figure 4 demonstrates that an MAE of 1.25 dB or better is needed for the AUCs of VF-MD/OCT-MD slopes to be above 0.80 for complete and partial substitution. When the MAE was 1.00 dB or better, the performance became nearly identical to VF-MD alone. For an MAE of 1.25 dB or better, 40% substitution had a higher AUC than simply using 40% fewer VFs. On the other hand, the AUC for 60% substitution only became higher than simply using 60% fewer VFs when the MAE was 1.50 dB or better.

**Figure 4:**
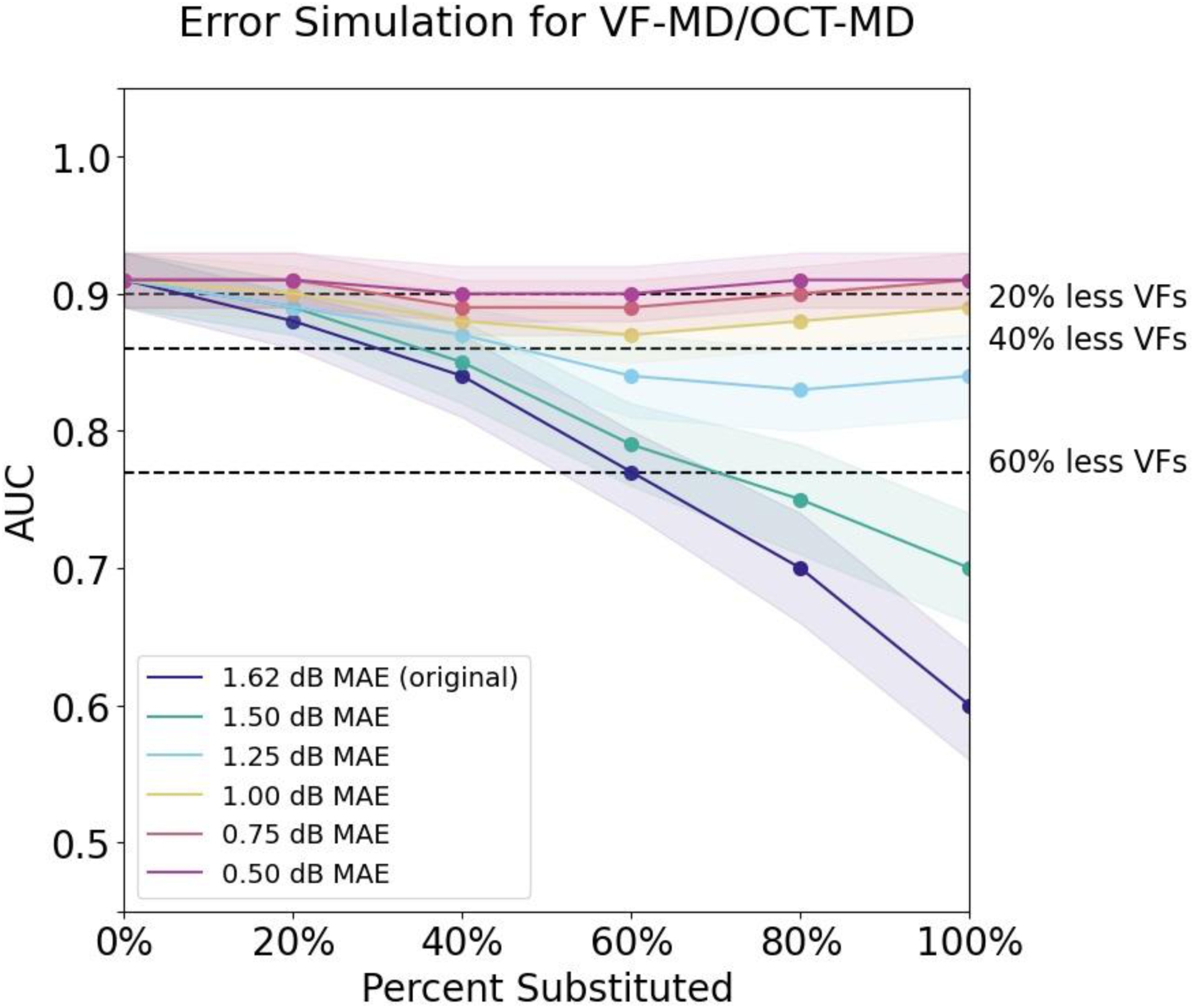
The predictive ability of VF-MD/OCT-MD slope with various levels of accuracy ranging from MAE of 0.50 to 1.50 dB. A substitution of 0% represents the performance of MD slopes calculated only from VF-MD (baseline VF-MD slope). A substitution of 100% represents the performance of MD slopes calculated from only OCT-MD. Dashed lines represent the performance of baseline VF-MD slopes using 20%, 40%, and 60% fewer VFs.

We performed a similar error simulation to evaluate the diagnostic ability of VF-MD + OCT-MD slopes calculated using OCT-MD estimates with an MAE of 1.50, 1.25, 1.00, 0.75, and 0.50 dB. We found that regardless of the accuracy of OCT-MD estimates, using OCT-MD as additional data points for MD slope calculation did not improve the AUC compared to using VF-MD alone.

### Sensitivity Analyses

In our first sensitivity analysis, we repeated the main analysis using only VF studies and OCT scans performed on the same day to address possible temporal bias introduced when estimating VFs from OCTs paired up to one year apart. The structure-function model had minimal improvement in performance, with an MAE of 1.59 dB for the OCT- MD estimates compared to an MAE of 1.62 dB for the OCT-MD estimates from the original model. We also achieved similar results to our original analysis when examining the predictive ability of the OCT-MD estimates to predict glaucoma progression.

However, it is important to note that the sample size for the progression dataset was substantially smaller with the adjusted inclusion criteria compared to the original inclusion criteria (n = 562 vs. n = 4,044).

In our second sensitivity analysis, we repeated the main analysis but required progressing eyes to have an MD slope worse than 0.50 dB/year, p-value < 0.05, and confidence intervals to be within +/- 0.25 dB of the calculated slope. Among the 4,044 eyes in the progression dataset, 19 were considered progressing using the above criteria. Similar trends were seen. The AUCs of partially substituted VF-MD/OCT-MD slopes were lower than the baseline VF-MD slope and were worse with increasing substitution percentages. VF-MD/OCT-MD slopes with 20% substitution had a similar AUC to the baseline VF-MD slope, but the same was true for VF-MD slopes using 20% fewer VFs. The AUCs of the VF-MD + OCT-MD slope were similar to the baseline VF-MD slope.

## Discussion

In this study, we developed a machine learning model trained on optic nerve head OCT measurements to estimate MD with an error (MAE) of 1.62 dB. The error of the OCT- MD estimates increased with increasing disease severity. Completely substituting VF-MD with OCT-MD resulted in significantly lower AUCs compared to using VF-MD alone. Partially substituting only 20% of VF-MD with OCT-MD had a statistically similar AUC compared to using VF-MD alone. However, partial substitution did not perform better than simply using 20% fewer VFs. Using OCT-MD as additional data points with VF-MD for MD slope calculation did not improve the AUC regardless of the MAE. OCT-MD estimates predicted progression as well as VF-MD alone when the MAE was at or below 1.00 dB.

### Model Performance for Estimating Mean Deviation

When predicting VF-MD from OCT data, our structure-function SVM model achieved an overall MAE of 1.62 dB. Prior classical machine-learning and deep-learning models were also trained with structured data, such as thickness measurements, and had mean errors ranging from 3 – 5 dB.^12,34,35^ Our MAE also compares well to more recent studies with sophisticated deep learning models incorporating unstructured data from unsegmented OCT images, which contain much more information than tabular RNFL thickness measurements (MAE ranging from 2.3 – 2.8 dB).^11,36,37^ Our findings are also consistent with those of Wong et al. (2022), who compared different machine learning models trained on global RNFL thickness measurements to estimate global VF-MD and found that gradient-boosted decision trees and SVM performed significantly better than other models, including some deep learning models.^35^

Several reasons may explain the better MAE observed in our study compared to previous work. Our structure-function model was trained on a substantially larger dataset (more than 50,000 OCT-VF pairs). It was trained on additional optic nerve features such as cup volume, disc area, rim area, and cup-to-disc ratio, not just RNFL thickness measurements. Unlike previous studies, we limited OCT-VF pairs used to train the model to only those that fall within the dynamic range of the OCT imaging instrument. The poor accuracy and variability of OCT-MD estimates as visual function worsens is well documented, and the reduced dynamic range of RNFL thicknesses in later stages of glaucoma is a frequently cited reason.^15,34,38^ Moreover, limiting our study population to the dynamic range also limits the generalizability of our model. The model was trained on mostly suspect or mild glaucoma, as eyes with more advanced disease would likely fall outside of the dynamic range. The VF measurements from more advanced diseases are known to be much more variable, which could explain the higher MAEs observed in prior studies.^9–14,16,39^

Since our study aimed to evaluate the feasibility of utilizing OCT-MD in trend-based analysis, we decided to use the conditions that would provide the most accurate estimations. Although this means the generalizability of our findings is likely limited to eyes with earlier stages of glaucoma, it is unlikely that including eyes outside the dynamic range, such as those with moderate or advanced glaucoma, would change the main findings of our study. If it is not possible to use OCT-MD to improve the ability to detect progression in eyes with earlier stages of disease, it is unlikely that OCT-MD would be helpful in eyes with later stages of disease where OCT-MD estimations would be even more inaccurate and variable.^39^

### Detecting Glaucoma Progression with OCT Estimated Mean Deviation

Combining OCT-MD with VF-MD, either through substitution or addition, did not significantly improve the ability to detect progression. Completely or partially substituting VF-MD with OCT-MD led to a worse ability to detect progression than using VF-MD alone. Although only substituting 20% of VF-MD led to statistically similar predictions compared to VF-MD alone (AUC of 0.86 vs 0.87), the same could be achieved by simply using 20% fewer VFs when calculating the rate of MD worsening. When using OCT-MD as additional data points to calculate the rate of MD worsening, the AUC was statistically similar to VF-MD alone, regardless of the MAE. However, when MAE was 1.00 dB or better, the OCT-MD estimates could be considered for substitution as the performance was similar to VF-MD alone.

If a low enough MAE could be achieved, there are multiple potential advantages to being able to convert an RNFL measurement to MD that we could expect. Since patients often alternate between OCT imaging and VF testing, one may detect glaucoma progression earlier since less time is needed to estimate the rate of MD worsening by using both OCT-MD and VF-MD. Being able to substitute VF-MD with OCT-MD in these situations offers great flexibility in the amount and type of testing needed to produce accurate trend assessments. Due to the variability of VF testing, especially for later stages of disease, at least 10 VFs are required to obtain the most accurate progression rate estimate.^40^ Adding OCT-MD as additional data points with VF-MD in these trend-based analyses may also allow one to obtain more accurate progression rate estimates in less time.

Despite the numerous efforts directed at developing structure-function models, there is currently no established evidence-based accuracy threshold that investigators can use to validate the clinical utility of their models. Our work demonstrates that OCT-MD estimates with margins of error greater than 1.00 dB did not provide clinical value in detecting glaucoma progression using trend-based analysis, even when combined with VF-MD measurements through partial substitution or addition. Figure 2 demonstrates that only when OCT-MD is used to predict VF-MD measurements within a 0 to –2 dB range the MAE could be better than 1.00 dB. For most clinical situations, detecting VF worsening within such a narrow range would be challenging. In a best-case scenario, current OCT-MD estimates may help detect worsening in pre-perimetric glaucoma.

More work is needed to produce OCT-MD estimates that are accurate enough to estimate progression rates. Several improvements can be made to our model to achieve the required accuracy. We only used mainly RNFL thickness measurements to predict MD, but more accurate estimations may be achieved if we develop a deep learning model that utilizes the RNFL thickness measurements and the corresponding raw, unsegmented OCT image. Lazaridis et al. (2022) previously demonstrated that an ensemble model utilizing the OCT image and the RNFL thickness profile had approximately a 22% lower MAE than a model using only the RNFL thickness profile.^37^ They suggested that OCT images contain additional information, such as vascular features that may be relevant to the structure-function relationship, as retinal ganglion cell loss in glaucomatous eyes with VF damage is associated with decreased regional retinal blood flow.^41,42^ Indeed, some studies have found that combining vasculature measurements from OCT angiography with structural OCT measurements can improve the ability to assess VF defects.^43,44^ Another important structural feature that can improve prediction accuracy is macular information. Yu et al. (2021) found that a deep learning model using both macular and optic nerve head scans had a lower median absolute error for predicting MD than either alone.^36^ Studies suggest that macular OCT also has a larger dynamic range than optic nerve head OCTs and may be more useful in predicting visual function in the later stages of glaucoma which can potentially help address the poor accuracy observed with structure-function models in later stages of disease and improve generalizability.^45–47^ Additionally, macular changes can be seen in early glaucoma, and incorporating this information could also further improve the estimations at earlier stages of disease.^48–50,50–54^

Another approach to detecting glaucoma progression using structure-function models that we did not investigate is predicting pointwise VF measurements. In addition to global VF indices, there are efforts to use OCT to estimate individual pointwise VF sensitivities.^10–12,15,34,37,55^ Predicting threshold sensitivities would maintain the spatial relationships between structural features that would not be reflected in an estimated summary metric such as MD. These spatial relationships may help in the early detection of localized disease progression before they can affect the global VF metrics. However, predicting pointwise measurements is a significantly more complex task than predicting global measurements.

Since the ultimate goal is to detect disease progression, other approaches that are important to continue investigating are models that predict progression directly rather than computing a VF metric from an OCT metric and then performing a subsequent analysis to predict disease progression. Models utilizing this approach have achieved good performance (AUC > 0.80) in predicting VF worsening from longitudinal OCT information.^33,56–58^ In our previous study, we used a gated-transformer network to predict VF worsening based on MD slope with longitudinal OCT scans.^33^ An advantage of this approach is that we could detect spatially dependent structural changes over time with our deep learning model, unlike this study. However, the model is limited by the fact that it requires a minimum of 5 OCT scans as an input (a disadvantage that is not present with the conversion approach). While waiting for enough tests, patients may experience additional vision loss or be lost to follow-up. Consequently, treatment decisions are often needed after only a few visits. These drawbacks also highlight the importance of models identifying future disease worsening with an early or limited diagnostic dataset.^58^

There are important limitations to acknowledge in our study. Although our SVM model achieved a low MAE, we only used tabular data from segmented RNFL measurements. As mentioned earlier, it is possible that if we incorporated the unsegmented OCT images, macular OCTs, or OCT angiography into a deep-learning model, we could achieve better MD estimates. We only investigated glaucoma worsening through MD slope but other trend-based analyses such as VFI slope may produce different results. Global VF indices also cannot show spatial relationships, and there is growing evidence that regional or sectoral changes demonstrate better agreement between structure and function.^8^ Predicting pointwise threshold sensitivities rather than global indices may be more useful for evaluating progression in other situations, such as with event-based analysis. When conducting our error simulations, we artificially reduced the residual for each estimate by a certain percentage to mimic a structure-function model with better accuracy, but this assumes that improvements can be made to the model that would improve the accuracy universally among all eyes with differing baseline characteristics and reduce error across all levels of VF damage in the same proportions. Additionally, our structure-function model is trained to predict a single paired VF taken at a single clinic visit and this assumes that the single VF is a true representation of the eye’s visual function. However, VF measurements are variable, especially at later stages of disease, and the error observed with the OCT-MD estimates for given measurement will be partly due to this inherent variability. Another approach could involve predicting an average of multiple VF tests as the ‘best available estimate’ of the true visual function.^37^

However, it would be impractical to obtain multiple VF measurements for each patient on a single visit and do so on a large scale to train a model. The VF studies included are obtained from a mix of testing strategies, and this could potentially confound the MD measurements, but this may only be a limitation for later stages of glaucoma. Prior studies have shown that SITA Faster and SITA Standard perform similarly in mild disease, and performance differences are more pronounced in later stages of disease.^59–63^ As the majority of our eyes are suspect or mild glaucoma (93%), we have less concern for measurement bias. Lastly, VF and OCT were paired within one year and this assumes no glaucomatous changes occurred during this timeframe. Although Chauhan et al. (2014) have shown that most eyes have slow rates of VF progression. It is unlikely that significant changes occurred between OCT scans and VF tests.^64^ Moreover, our sensitivity analysis utilizing only OCTs and VFs obtained on the same day did not change our results, albeit it could be underpowered.

In conclusion, we developed a machine learning model to estimate MD from optic nerve head OCT scans with a low prediction error compared to other structure-function models in the literature. We used the model to estimate VFs from paired OCTs in patients with longitudinal data. We found that even if OCT-MD is combined with VF-MD, either through substitution or addition, it did not improve the ability to detect disease progression compared to VF-MD alone. We conducted an error simulation analysis to determine the accuracy needed for our model to detect glaucoma progression. We found that when the MAE is 1 dB or better, OCT-MD could substitute VF-MD in trend-based analysis. Future work developing structure-function models should aim to achieve this lower level of prediction error to ensure the clinical utility of such models to detect functional change over time.

## Data Availability

All data produced in the present study are available upon reasonable request to the authors

## Abbreviations and Acronyms

VF: visual field
MD: mean deviation
dB: decibels
OCT: optical coherence tomography
RNFL: retinal nerve fiber layer
ML: machine learning
MAE: mean absolute error
SVM: support vector machine
AUC: area under the curve

